# How Healthcare Congestion Increases Covid-19 Mortality: Evidence from Lombardy, Italy*

**DOI:** 10.1101/2020.10.27.20221085

**Authors:** Gabriele Ciminelli, Sílvia Garcia-Mandicó

**Author notes:** The views expressed in this paper are those of the authors and do not represent those of the OECD or its member countries. We are grateful to Richard Baldwin, Travers Barclay Child, Emanuele Ciani, Viktar Fedaseyeu, Pilar Garcia-Gomez, Massimo Giuliodori, Sergi Jiménez-Martín, Luca Marcolin, Owen O’Donnell, Magdalena Rola-Janicka, Stefan Thewissen and seminar participants at CEIBS and the OECD for helpful feedback and discussion. Ciminelli: Asia School of Business, MIT Sloan School of Management; Garcia Mandicó: OECD.

## Abstract

**Background:** The Covid-19 pandemic has caused generous and well-developed healthcare systems to collapse. This paper quantifies how much system congestion may have increased mortality rates, using distance to the ICU as a proxy for access to emergency care.

**Methods:** We match daily death registry data for almost 1,500 municipalities in Lombardy, Italy, to data on geographical location of all ICU beds in the region. We then analyze how system congestion increases mortality in municipalities that are far from the ICU through a differences-in-differences regression model.

**Findings:** We find that Covid-19 mortality is up to 60% higher in the average municipality – which is 15 minutes driving away from the closest ICU – than in a municipality with an ICU in town. This difference is larger in areas and in days characterized by an abnormal number of calls to the emergency line.

**Interpretation:** We interpret these results as suggesting that a sudden surge of critical patients may have congested the healthcare system, forcing emergency medical services to prioritize patients in the most proximate communities in order to maximize the number of lives saved. Through some back-of-the-envelope calculations, we estimate that Lombardy’s death toll from the first Covid-19 outbreak could have been 25% lower had all municipalities had ready access to the ICU. Drawing a lesson from Lombardy’s tale, governments should strengthen the emergency care response and palliate geographical inequalities to ensure that everyone in need can receive critical care on time during new outbreaks.

**Funding:** No funding.

## 1 Introduction

The first wave of Covid-19 strained healthcare systems in many countries, abounding a series of lessons for governments to prepare for new outbreaks. The principal lesson has highlighted the need to strengthening hospitals and intensive care units (ICUs) capacity. As such, policies have so far mostly focused on expanding already existing emergency infrastructure and building field hospitals near existing medical centres.^1^ While necessary, this does little to reduce geographical inequalities in emergency care coverage. This paper highlights the need of tackling such disparities by quantifying the mortality effects of having communities underserved by emergency care when the system is under severe strain.

The analysis focuses on Lombardy, which offers a good case to study. The region is one of Italy’s wealthiest and is also renowned for having one of the best healthcare infrastructures in the country, which itself has fairly high rates of ICUs per capita. ^2^ Yet, Lombardy suffered extremely high mortality rates during its first Covid-19 outbreak, which cannot be explained by traditional epidemiological models, such as SIR and SEIR.^3,4,5^ These models feature an exogenous probability of dying once individuals become infected. However, as noted by Favero, if the healthcare system is saturated and infected people cannot access to the ICU, such probability increases, becoming endogenous to the level of system congestion. ^6^

This paper analyzes how much system congestion may have contributed to the high mortality rates observed during Lombardy’s first Covid-19 outbreak, using distance to the ICU as a proxy for access to emergency care. To carry out the analysis, we match highly granular daily death registry data for almost 1,500 municipalities to information on geographical location and number of all ICU beds across Lombardy. We complement the dataset with daily data on the volume of calls to the emergency line, as well as data on a number of co-factors of Covid-19 mortality.

We start by showing that, despite its generosity, the region’s healthcare system is characterised by significant disparities: the average distance in minutes of driving to the nearest ICU is three times as large for municipalities in the mountainous subregion of the Alps as for those in the metropolitan area of Milan. This is important because, when the system is overwhelmed and there is not enough time to attend everyone in need, emergency medical services may have to prioritize patients in the most proximate communities, at the expense of reducing geographical coverage, in order to maximize the number of lives saved. ^7^ To test whether distance to the ICU has any effect on Covid-19 mortality, we develop a differences-in-differences regression model. We find that mortality is up to 60% higher in the average municipality – which is 15 minutes of driving away from the closest ICU – than in a municipality with an ICU in town.

Of course, distance on its own does not imply that critical patients cannot get to the ICU on time. After all, some patients require transportation from remote communities to the ICU also in normal times, and usually they get there on time. Distance to the ICU only becomes a determining factor when the burden on the emergency care system is high, as capacity to serve everyone in need is reduced. We proxy for system congestion using data on calls to the emergency line and find that the additional effects of Covid-19 on mortality in municipalities that are farther away from the ICU is stronger in days and areas characterized by an abnormal volume of calls to the emergency line, pointing to system congestion as a plausible explanation.

We then quantify how much system congestion may explain the high mortality rates observed in Lombardy’s first Covid-19 outbreak. To do so, we perform some back-of-the-envelope calculations to compare the number of deaths that occurred in actuality to those that would have occurred in a hypothetical scenario in which all communities had an ICU in town. We find that Covid-19 deaths would have been about 25% less in such as a scenario, meaning that many lives could have been saved through more widespread critical care coverage. The rest of the paper is organized as follows. We close Section 1 by putting our analysis in context. Section 2 describes the dataset and methodology. Section 3 presents the results and Section 4 concludes.

### Research in context

#### Evidence before this study

Epidemiological models such as SIR and SEIR are at the center of a quickly growing literature assessing how mitigation policies can be optimally set to minimize the burden on the economy while reducing the number of fatalities. However, these models fail to predict the mortality observed in outbreak epicenters, where healthcare systems, and in particular emergency care systems, are overwhelmed. Another strand of the literature seeks to understand the causes behind the severity of Italy’s first Covid-19 outbreak.

#### Added value of this study

We contribute to these two strands of work by focusing on the congestion of the healthcare system as an important reason behind the high death toll of Lombardy’s first Covid-19 outbreak, which cannot be explained by existing epidemiological models. To our knowledge, this is the first study to quantify how an uneven distribution of ICUs across communities may increase Covid-19 mortality when the health system is overburdened. Our results – which are robust to controlling for a host of co-factors of Covid-19 mortality – indicate that geographical differences in healthcare coverage, together with overwhelmed healthcare systems, account for a significant fraction of the observed Covid-19 mortality in the region.

#### Implications of all the available evidence

Drawing a lesson from Lombardy’s tale, governments around the world should invest in strengthening their emergency care response, and in particular, devising measures to palliate geographical inequalities in emergency care coverage in order to ensure that everyone in need can receive critical care on time. They should improve pre-hospital emergency services, by clarifying the first point of contact for possible Covid-19 cases, expanding capacity to manage large volumes of calls, and improving phone triage to better prioritize care delivery. They should also invest in building ambulance capacity and, ideally, mobilize ICUs more evenly across the territory. All these factors are key to help reducing mortality in the new waves of Covid-19.

## 2 Methods

### Data

We source death registry and census population data at the municipality-level from ISTAT, the Italian Statistical Agency. ^8,9^ Census data provide information on the resident population as of January/1^st^, while death registry data provide information on daily deaths for the January/1^st^ to May/15^th^ period for the years 2015 to 2020, for almost all Italian municipalities. To measure Covid-19 mortality, we rely on the concept of excess deaths – that is, the difference between deaths for all causes during the Covid-19 epidemic and deaths that would be expected under normal circumstances. We prefer this approach over using official Covid-19 fatality data because these vastly undercount the real number of Covid-19 deaths, as we show in a companion paper. ^10^ Moreover, focusing on excess deaths has the key advantage that underlining data are much more granular than official fatality data, which is crucial for our identification strategy, as it will become clear below.

Our focus is on the Lombardy region, which is universally considered as Europe’s ground zero for Covid-19 and the epicenter of Italy’s first outbreak, making up for about 50% of all fatalities, with less than 17% of the overall population. The data cover 1,455 municipalities, together accounting for almost 98% of Lombardy’s population.

Lombardy’s emergency medical services are organized in four geographical subregions. Municipalities in the mountainous area belong to the Alps subregion, those in the lakes area are part of the Lakes subregion, those in the flatlands belong to the Po Valley subregion, while municipalities around Milan constitute the Metropolitan subregion. To measure congestion in the healthcare system, we follow van Dijk and co-authors and source data on the volume of daily calls to the emergency line, by subregion and reason for the call.^11^ These data are taken from *L’Eco di Bergamo*.^12^

We also source data on location and areas of specialization of each hospital and private clinic from Lombardy’s institutional database.^13^ We use this information to construct a municipality-level variable measuring distance to the nearest ICU. If there is an ICU in town, we set it to zero. Otherwise, the variable measures the distance, in minutes of driving, to the nearest municipality with an ICU (taken from ISTAT).^14^ Admittedly, ICU capacity has been strengthened during the Covid-19 epidemic and these additional emergency ICU beds are not recorded in our data on health facilities, which provide a snapshot as of end-2019. However, this does not impact our distance-to-ICU measure because capacity expansion was concentrated in municipalities that already had ICUs (mostly in Bergamo, Crema, and Milan).

The dataset is complemented with variables capturing slow-moving socio-demographic, labor market and territorial characteristics that we use to control for potential co-factors of Covid-19 mortality. As most of these variables are not available at a regular frequency, we compute their means over the 2015-2019 period and treat them as time-invariant factors. Appendix Table A2 provide detailed information on their sources and coverage. Next, we discuss a few stylized facts emerging from the data.

### Daily deaths increased four-fold in less than a month

Before the detection of the first community case, deaths in 2020 match very closely deaths in 2016 (see Appendix Figure A1). The severe effects of Covid-19 on mortality is underscored by the exponential increase in deaths following the detection of the first community case – at their peak, roughly a month after detection, deaths in 2020 are about four times as large as deaths in 2016. Also worth noting, excess mortality reaches particularly high levels in the Alps and the Po Valley subregions (see Appendix Figure A2). Overall, the virus may have contributed to the death of up to 0.4% of the local population in the Alps, and about 0.3% in the Po Valley.

### An ambulance roundtrip to the ICU takes 30 minutes on average

We then move on to the emergency care system. Figure 1 below characterizes municipalities according to distance to the closest ICU. Only less than 4% of all municipalities have an ICU in town. As expected, these tend to be the larger municipalities. However, together they only account for less than a quarter of Lombardy’s population.

**Figure 1:**
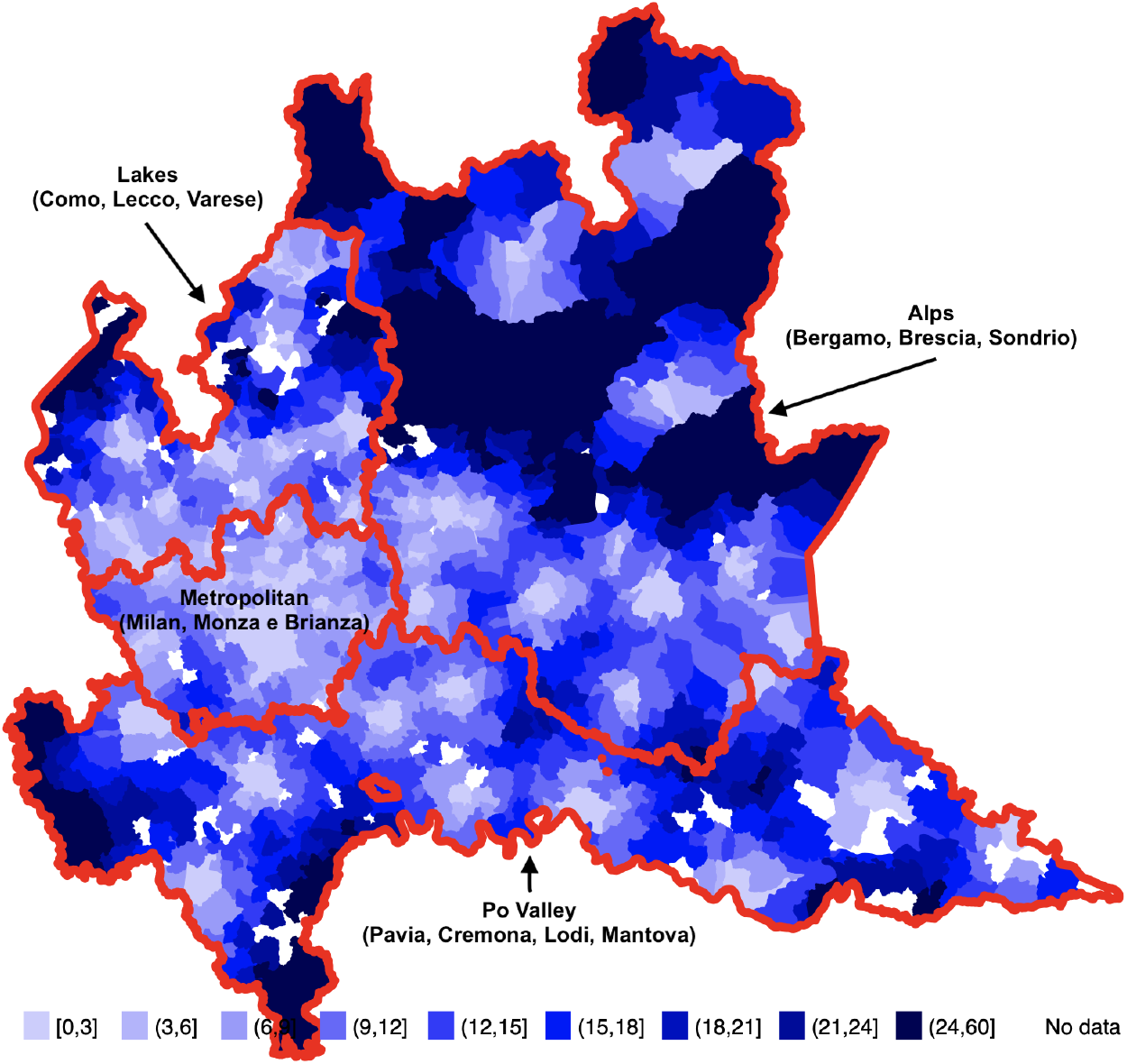
Distance to closest ICU in minutes of driving. *Notes:* This map characterizes municipalities by distance to the closest ICU using different shades of blue. Distance is calculated in minutes of driving (one-way trip). Lighter (darker) colors denote smaller (larger) distances. Red lines denote the borders of the four different geographical areas under which the emergency care service is organized.

The average municipality is 15 minutes away from the closest one with an ICU in town. But this figure masks large disparities between the subregions. Mean distance to the ICU is reduced to just 7.5 minutes for municipalities in the Metropolitan subregion, while about a quarter of all municipalities in the Alps subregion are further than 25 minutes away, meaning that an ambulance roundtrip may take more than an hour.

### Calls to the emergency line more than doubled in the Alps subregion

Figure 2 below focuses on calls to the emergency line, distinguishing between total calls and calls for respiratory or infectious disease reasons (respiratory reasons thereafter).

**Figure 2:**
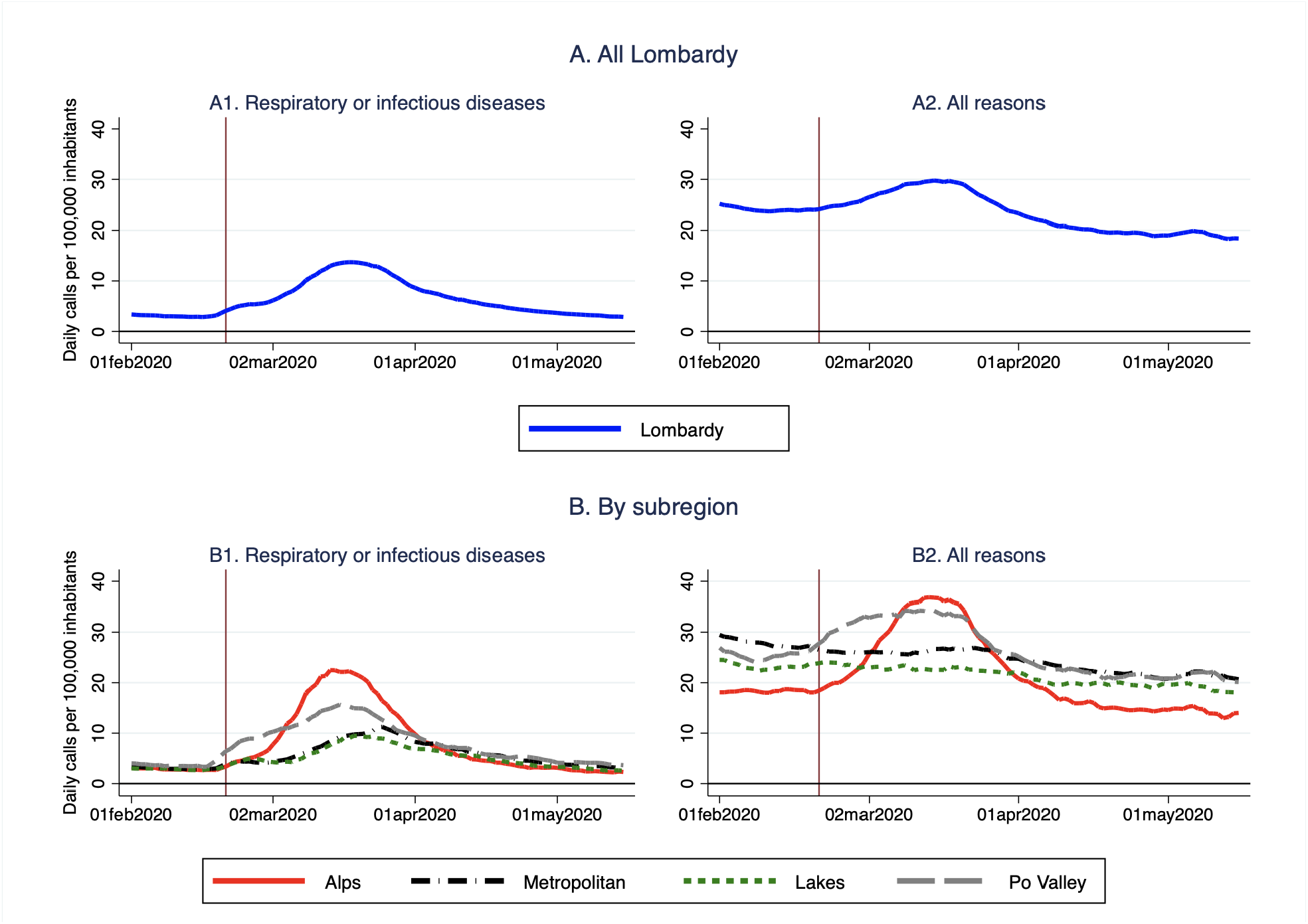
Calls to the emergency line. *Notes:* The figure shows the volume of daily calls to the emergency line per 100,000 inhabitants, for respiratory or infectious disease reasons (Panel A) and for all reasons (Panel B), in the Lombardy region.

Overall in Lombardy, calls for respiratory reasons surge more than four-fold in the month following the detection of the first community case (denoted by the vertical line), from about 3 to almost 14 per 100,000. After peaking, they decrease very slowly, returning to pre-Covid-19 levels only three months following detection (Panel A1). The increase in total calls (calls for any reason) is more nuanced and the reversal to pre-Covid-19 levels happens much earlier, likely due to seasonal factors and the fact that the lockdown imposed by the government to contain the epidemic reduced road and workplace accidents (Panel A2). Panels B1 and B2 zoom in on calls received in each of the four subregions. While in the Alps calls for respiratory reasons surge more than 8-fold and total calls double, the increase in calls for respiratory reasons is less than 5-fold in the Po Valley and less than 4-fold in the Metropolitan and the Lakes subregions. In the latter two the total volume of calls barely increase.

All in all, this visual inspection of the data suggests that, although calls for respiratory reasons increased across the board, the Alps subregion – which is also the one with the most uneven distribution of ICUs – may have particularly struggled to cope with the surge in demand for critical care. In what follows we illustrate the methodology used to formally analyze the insights that emerged from this first look of the data.

### Empirical Methodology

We first quantify the effect of Covid-19 on the mortality rate. To do so, we follow closely the methodology that we developed in two companion papers. ^10,15^ Specifically, we rely on a differences-in-differences approach to estimate the dynamic effects on the mortality rate, using the year 2016 as counterfactual of what mortality would have been in absence of Covid-19. The choice of using the year 2016 follows from a visual inspection of the data (see Appendix Figure A1), but the results are robust to using mean 2015-2019 mortality as alternative counterfactual. The equation that we estimate is as follows:

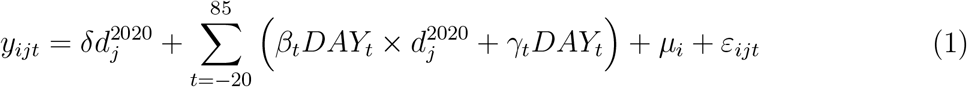

where *y*_*ijt*_ measures daily deaths per 100,000 inhabitants in municipality *i*, at within-year time *t*, for year 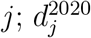 is a dummy variable taking value equal to 1 in 2020 and 0 otherwise; *DAY*_*t*_ are within-year time effects, taking value 1 in each particular day of the year and 0 otherwise; *µ*_*i*_ are municipality fixed effects; and *ε*_*ijt*_ is an idiosyncratic error, clustered at the municipality-level. The summation term ranges from −20 to +85 since we normalize the within-year time dimension *t* so that it takes value equal to 0 on the day in which the first community case was detected (February/21^st^) and negative (positive) values on days before (after). For the estimation, we use the least squares method with population analytical weights.

Appendix Figure A3 depicts the effect of Covid-19 on mortality, obtained plotting the 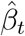 coefficients estimated from Equation (1). The effect peaks exactly a month after onset, at slightly below 8 deaths per day per 100,000 inhabitants and then slowly decreases, until becoming statistically insignificant 44 days after the peak. These estimates are robust to using average deaths in the five preceding years (2015 to 2019) as counterfactual (also reported in Appendix Figure A3).

Next, we turn to the congestion of the emergency care system as one potential factor that may have increased mortality. We start by uncovering a positive relationship between daily mortality rates and the variable measuring distance to ICU (see Appendix Figure A4). More precisely, being 10 minutes farther away from the ICU is associated with about 1 more death per 100,000 inhabitants per day, on average, during the entire epidemic period. This relationship is highly statistically significant.

To more formally estimate the effects of the uneven distribution of ICUs on mortality, we extend Equation (1) by adding an interaction term between the within-year effects, the year 2020 dummy and our variable measuring distance to the ICU. More specifically, the equation that we estimate is as follows:

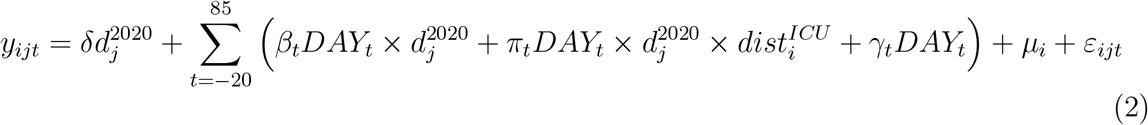

where 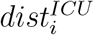 is the variable measuring distance, in municipality *i*, to the closest municipality with an ICU in town; and the rest of the notation is as before. As for Equation (1) above, the estimation is through OLS with standard errors clustered at the municipality-level.

## 3 Results

### Up to 60% higher mortality in municipalities 15 minutes away from the ICU

Figure 3 below shows the results. Precisely, it compares the mortality effect of Covid-19 in the average municipality, in which the closest ICU is 15 minutes driving away (red line with crosses, given by 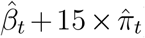), to the same effect in municipalities with an ICU in town (blue line, given by 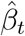). Shaded areas represent 95% confidence intervals. The figure also plots the total volume of daily calls to the emergency line (dashed black line, right axis).

**Figure 3:**
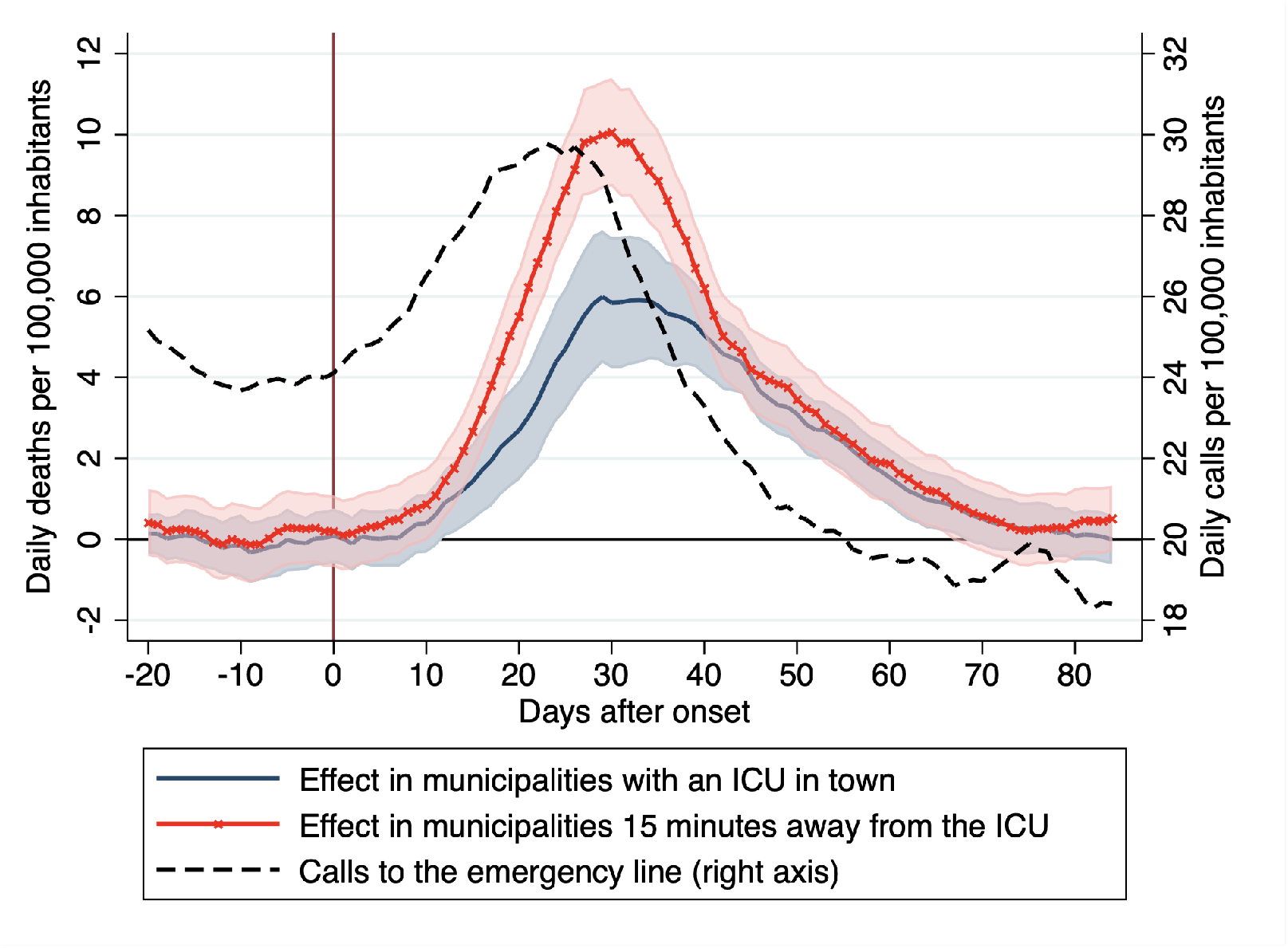
Effect of Covid-19 on the mortality rate and calls to the emergency line. *Notes:* The figure shows the dynamic effects of Covid-19 on in municipalities with an ICU in town (blue solid line) and in those where the ICU is 15 minutes driving away (red line with crosses) and compares these effects with the evolution of calls to the emergency line (black dashed line, right axis). Estimates are given by 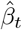 (municipalities with an ICU in town) and 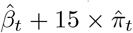 (municipalities with the closest ICU being 15 minutes driving away), obtained estimating Equation (2). Shaded areas represents 95% confidence intervals. The y-axes report daily deaths and calls to the emergency line per 100,000 inhabitants (left and right axes respectively). The x-axis reports the number of days since the detection of the first community case.

The average municipality experiences significantly higher mortality rates than those with an ICU in town at the height of the epidemic. At peak, the effect of Covid-19 on the mortality rate reaches 6 deaths per day per 100,000 inhabitants in municipalities with an ICU in town, while the same effect is about 10 deaths per day per 100,000 inhabitants, or over 60% higher, in the average municipality, which is 15 minutes away from the ICU. The divergence in the mortality effect of Covid-19 across these two groups of municipalities match very closely the evolution of calls to the emergency line. Particularly, the additional effect in municipalities that do not have an ICU in town starts decreasing shortly after that the volume of calls to the emergency care eases up, and it becomes statistically insignificant once emergency calls return to pre-Covid-19 levels.

How can we explain the result that Covid-19 mortality rates are higher in communities that are more distant from the intensive care? One possibility is that the emergency care system struggled to cope with a surge in demand. Indeed, Sorbi reports that waiting times for emergency transportation swelled: to make a trip that usually took only 8 minutes, ambulances were taking an hour, and in some cases, they were not getting in on time.^16^ This congestion may have forced emergency medical services to prioritize serving more patients at the expense of reducing geographical coverage (see the report led by Vergano for evidence on the existence of prioritization guidelines).^7^

This begs the question of how many deaths could have been prevented if all communities were readily served by an ICU. To answer this question, we use the coefficients estimated in Equation (2) and perform some back-of-the-envelope calculations. First, we calculate the overall number of deaths that can be ascribed to Covid-19 in more than 26,000. Then we calculate the number of Covid-19 deaths in a hypothetical counterfactual scenario in which every municipality had an ICU in town. We find that deaths would have been slightly less than 20,500 in this counterfactual scenario. This means that about 28% of all Lombardy’s deaths in the first Covid-19 outbreak could have been prevented through better ICU coverage.

Before proceeding further, we check that our results are robust to controlling for other municipality characteristics that may correlate with distance to the ICU and that could also have an effect on mortality, such as population density, income, education, the demographic structure, the share of people employed in the healthcare sector, the number of ICU beds per capita, the share of people employed in essential sectors, distance from the epicenter and others. We also verify that our results are not driven by outliers and exclude municipalities with abnormal mortality rates and those in which the closest ICU is very far away. Finally, we check that our estimates are robust to using an alternative variable measuring distance to the ICU, in kilometers rather than minutes (taken from ISTAT).^14^ All the results from these robustness specifications are shown in Appendix Figure A5. For simplicity, the figure shows the additional effects of being 15 minutes away from the ICU, given by the 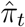 coefficients estimated from Equation (2), and compares it with the same effects estimated from the alternative specifications discussed above. Overall, the new estimates are very close to, and not statistically different from, our baseline, thus confirming the validity of our results.

### Larger effects when and where the emergency care system is congested

Next, we zoom in on the Alps subregion, where calls to the emergency line for respiratory reasons increased more than 8-fold and total calls more than doubled (see Figure 2 above), putting particularly high pressure on the emergency care system. If indeed distance to the ICU increases mortality when the system is congested, we should find larger effects in the Alps than in the other subregions, in which the increase in emergency calls was less pronounced.

To formally test this hypothesis, we twist Equation (2) to estimate two distinct sets of coefficients measuring the additional effect of distance to the ICU on mortality, one for the Alps subregion and the other for the rest of Lombardy. Figure 4 below shows these newly estimated coefficients. For simplicity, the figure only reports the additional effect of Covid-19 on mortality in municipalities that are 15 minutes away from the ICU relative to those with an ICU in town.

**Figure 4:**
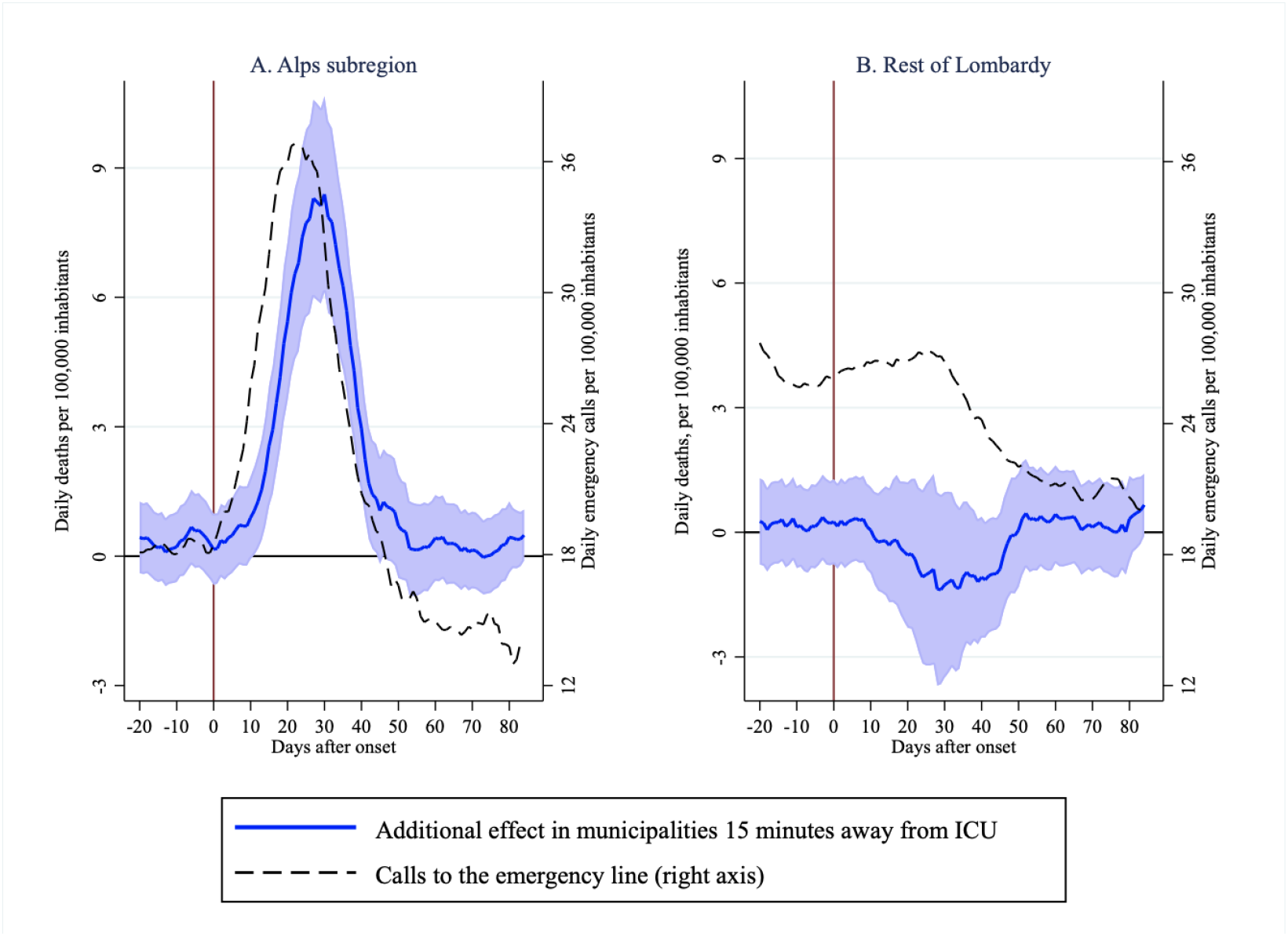
Additional mortality effects of being 15 minutes away from the ICU. *Notes:* The figure shows the additional effects of Covid-19 on mortality in municipalities that are 15 minutes away from the ICU relative to those with an ICU in town, distinguishing between municipalities in the Alps subregion (Panel A) and those in the rest of Lombardy (Panel B), and comparing this additional effect with the evolution of calls to the emergency line. Estimates are obtained estimating an expanded specification of Equation (2), which allows for a differential effect of distance to ICU on mortality in the Alps subregion vs. the rest of Lombardy. Blue solid lines denote the estimated effect, shaded areas are 95% confidence intervals, while dashed black lines depict the daily volume of calls to emergency system. The y-axes report daily deaths and calls to the emergency line per 100,000 inhabitants (left and right axes respectively). The x-axis reports days after onset.

The extra effect of being distant from the ICU on Covid-19 mortality is concentrated in the Alps subregion. There, municipalities that are 15 minutes away from the ICU experience up to 8 more deaths per day per 100,000 inhabitants. For the rest of Lombardy, instead, we do not estimate any statistically significant difference in mortality between municipalities that are far from the ICU and those with an ICU in town.

Looking at the volume of calls to the emergency line, we note that the dynamics of the extra effect of being distant from the ICU closely follow the evolution of emergency calls in the Alps, reinforcing the interpretation that system congestion may have prevented the transportation of critically ill patients to the emergency room on time. On the other hand, we observe that calls to the emergency line only slightly increase in the rest of Lombardy, which suggests that the system did not become congested there and helps rationalizing why more remote communities did not experience higher mortality rates there.

## 4 Discussion

We analyzed how emergency care congestion may have contributed to the high mortality rates observed in Lombardy during its first Covid-19 outbreak, using distance to the ICU as a proxy for access to critical care. We found that Covid-19-induced mortality was much higher in communities underserved by intensive care. Using the estimated coefficients we performed some back-of-envelope calculations to calculate how many deaths can be ascribed to system congestion. We found that more than 25% of all fatalities of Lombardy’s first outbreak may have resulted from system congestion.

Our results suggest that many Covid-19 deaths may have been prevented through better preparedness. Drawing a lesson from Italy’s tale, governments around the world should invest in strengthening their emergency care response. They should improve pre-hospital emergency services, by clarifying the first point of contact for possible Covid-19 cases, expanding capacity to manage large volumes of calls, and improving phone triage to better prioritize care delivery. They should also invest in building ambulance capacity, and, ideally, mobilizing ICUs more evenly across the territory. All these factors are essential to help reduce mortality during new outbreaks.

## Data Availability

Data available upon request

## Appendix

**Table A1:** Coverage.

| Subregion | mun. | % of all mun. | pop. (in 1,000) | % of all pop. |
| --- | --- | --- | --- | --- |
| Alps | 520 | 99.24 | 2759.545 | 99.61 |
| Metropolitan | 181 | 95.77 | 4165.865 | 98.52 |
| Lakes | 351 | 95.12 | 1908.318 | 97.22 |
| Po Valley | 403 | 96.41 | 1664.997 | 97.71 |
| Total | 1455 | 97.00 | 10498725 | 98.43 |

*Notes:* the columns "mun." and "% of all mun." report the number of municipalities covered and their share over all municipalities in the subregion. The columns "pop (in 1,000)" and "% of all pop." report the population covered (in thousands) and its share over all the regional population. The Alps subregion includes municipalities in the Bergamo, Brescia and Sondrio provinces. The Metropolitan subregion includes municipalities in the Milan and Monza e Brianza provinces. The Lakes subregion includes those in the Lecco, Como and Varese provinces, while the Po Valley subregion includes municipalities in the Pavia, Lodi, Cremona and Mantova provinces. Population for 2020 is imputed using 2019 growth rates.

**Table A2:** Variable descriptive, sources and coverage.

| Variable | obs. | mean | s.d. | min | max | source | definition |
| --- | --- | --- | --- | --- | --- | --- | --- |
| Distance to ICU, minutes | 1441 | 14.23 | 9.18 | 0.00 | 56.64 | <sup>13,14</sup> and own calculations | Distance to the closest municipality with an ICU, in minutes of driving |
| Distance to ICU, kilometers | 1441 | 14.12 | 9.91 | 0.00 | 56.89 | <sup>13,14</sup> and own calculations | Distance to the closest municipality with an ICU, in kilometers |
| ICU beds | 1455 | 0.07 | 0.41 | 0.00 | 7.00 | <sup>13</sup> | ICU beds, as % of population |
| Nursing home beds | 1455 | 0.65 | 1.60 | 0.00 | 20.79 | <sup>17</sup> | Beds in nursing homes, as % of population |
| Share of 80+ | 1455 | 6.65 | 2.31 | 1.15 | 23.34 | <sup>9</sup> | Share of people 80 years old and older, % of population |
| Share women | 1455 | 48.88 | 1.80 | 28.53 | 62.11 | <sup>9</sup> | Share of women in 18-65 population, % of population |
| Share graduates | 1455 | 53.7 | 8.87 | 18.37 | 85.38 | <sup>18</sup> | Share of high school graduates in 18-65 population, % of population |
| Mean income | 1452 | 14.18 | 2.37 | 3.03 | 30.70 | <sup>9,19</sup> and own calculations | Mean taxable income in 1,000 of euros |
| Commuting index | 1455 | 27.10 | 12.62 | 0.00 | 81.24 | <sup>19</sup> | External commuting index |
| Population density | 1455 | 2.54 | 1.09 | 0.33 | 9.43 | <sup>19</sup> and own calculations | 1,000 of people per squared kilometer of inhabited land |
| PM10 | 1455 | 60.85 | 18.04 | 23.33 | 81.67 | <sup>20</sup> | Number of days PM10 above limit, provincial level |
| Business shut-downs | 1455 | 52.17 | 13.22 | 8.50 | 92.57 | <sup>15</sup> | Share of employment in non-essential sectors, % total employment |

**Figure A1:**
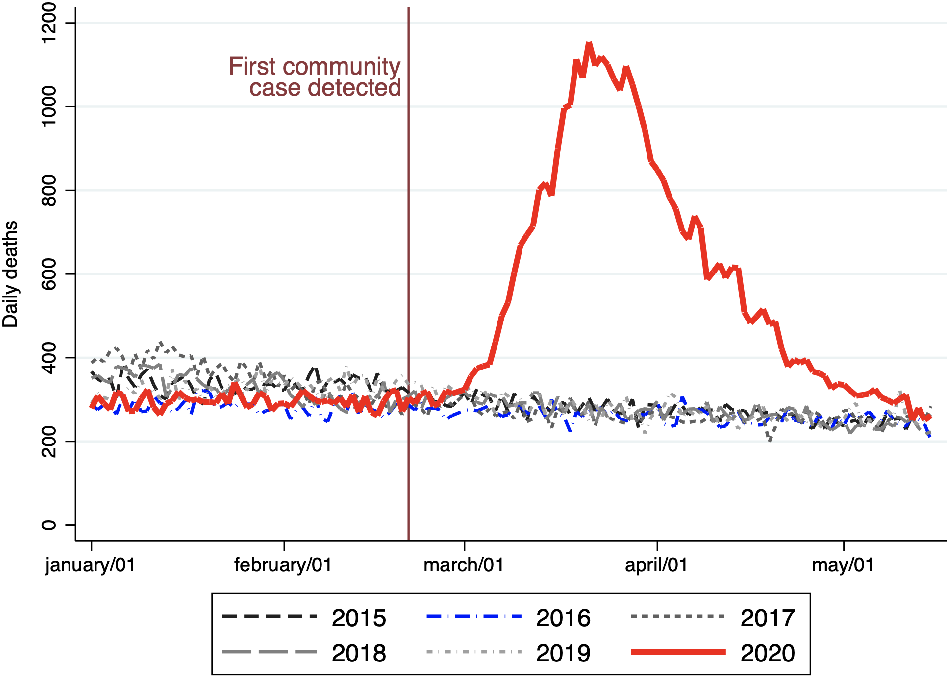
Deaths in 2020 compared to the five previous years. *Notes:* The figure compares daily deaths in 2020 (red solid line) to daily deaths in each of the five preceding years. The blue dashed line denotes deaths in 2016, which we use as counterfactual for the estimation of the effects of COVID-19 on mortality (see Section 2). The vertical maroon line denotes the day in which the first COVID-19 community case was detected, on February/21^th^).

**Figure A2:**
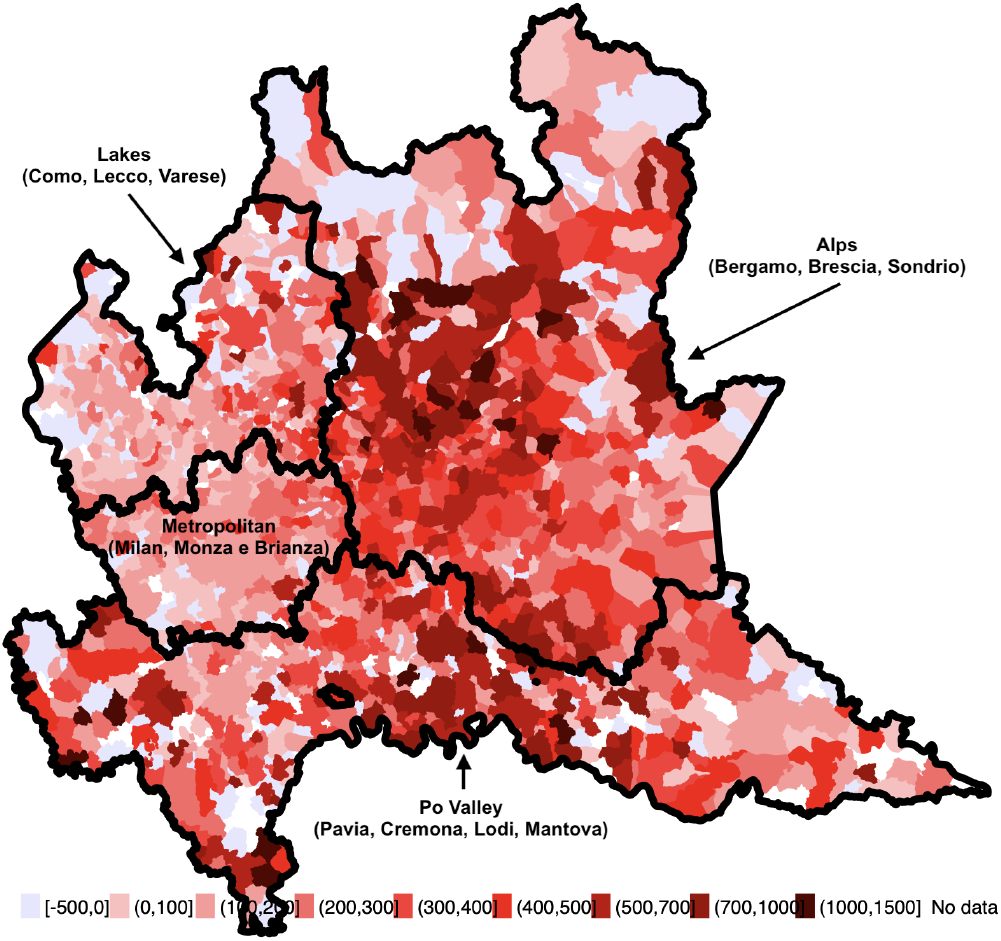
Excess mortality rate by municipality. *Notes:* This map characterises excess mortality rates, calculated per 100,000 inhabitants. Excess mortality rates are calculated by subtracting 2016 mortality rates to that in 2020. This method is akin to the empirical method we use for counterfactual estimation of the effects of COVID-19 on mortality (see Section **??**). Lighter (darker) colors denote lower (higher) excess mortality rates associated to COVID-19. The black lines denote the four different geographical areas under which the emergency care service is organized.

**Figure A3:**
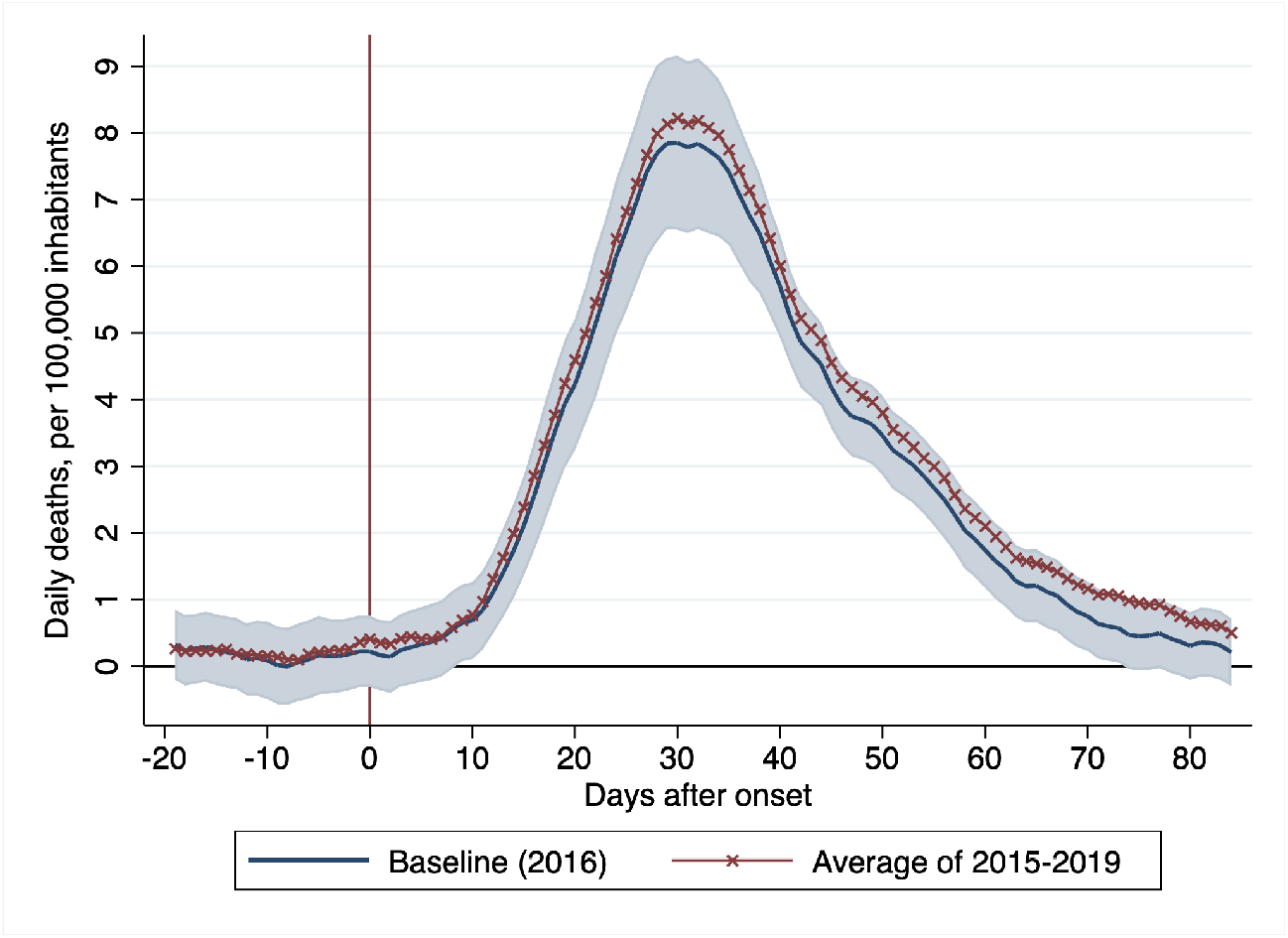
The dynamic effects of Covid-19 on the mortality rate. *Notes:* The figure shows the effect of Covid-19 on the mortality rate, measured in daily deaths per 100,000 inhabitants. Coefficients are estimated from Equation (1). The blue solid line show coefficients estimated using 2016 as counterfactual, while the blue shaded area depicts 95% confidence interval. The red line with crosses depicts estimates obtained using 2015-2019 mean mortality as counterfactual.

**Figure A4:**
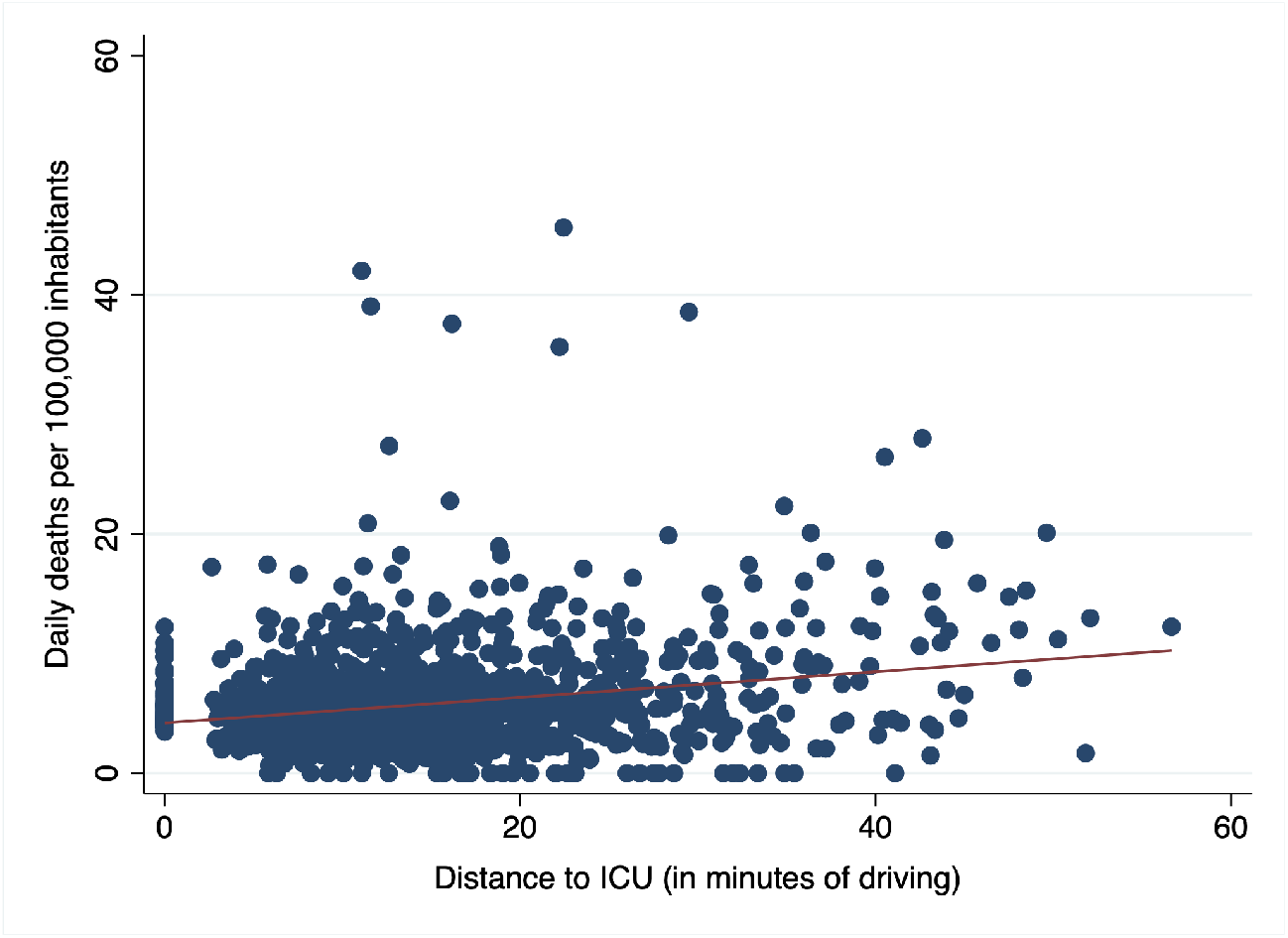
Distance to ICU and mortality rates. *Notes:* The figure depicts the relationship between distance to ICU and observed mortality rates. Precisely, it plots the average daily mortality rate per 100,000 inhabitants over the February/21/2020-May/15/2020 period onto distance to ICU, measured in minutes of driving for all municipalities in Lombardy.

**Figure A5:**
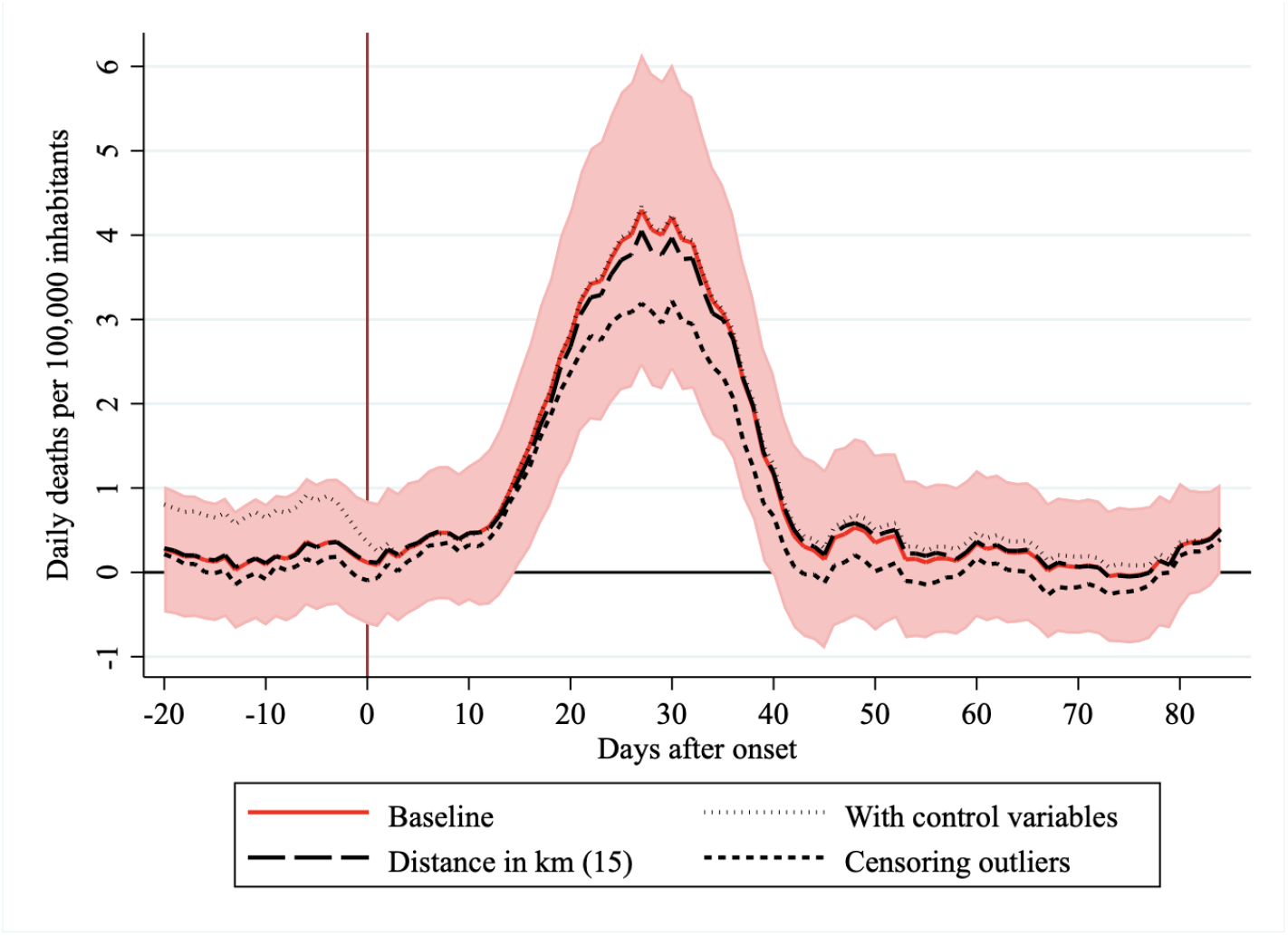
Robustness checks on baseline estimates. *Notes:* The figure shows the additional effect of Covid-19 on the mortality rate in municipalities that are 15 minutes driving away from the closest ICU relative to the effect in municipalities with an ICU in town. The red solid line depicts our baseline estimates (15 × *π*_*t*_ from Equation (2)). The dotted black line depicts estimates obtained augmenting Equation (2) by adding a set of control variables: share of women in working age population, share of high school graduates in working age population, mean income, share of 80-plus in population, population density, mean income, external commuting index, number of days in which PM10 is above limit, per-capita ICU beds, per-capita nursing home beds and distance to the outbreak epicenter. The long-dash line depicts estimates obtained using a different distance to ICU variable, in kilometeres rather than minutes of driving. The short-dash line depicts estimates obtained by censoring observations with outlier mortality rates or distance to the ICU. The y-axis measures daily deaths per 100,000 inhabitants, the x-axis measures days since the detection of the first community cases (denoted by a vertical maroon line).

## References

[1] Lionel Laurent. Are hospitals ready for covid’s second wave? Bloomberg Opinion, Oct 2020.

[2] Niel McCarthy. The countries with the most critical care beds per capita, 2020.

[3] Andrew Atkeson. What will be the economic impact of covid-19 in the us? rough estimates of disease scenarios. Working Paper 26867, National Bureau of Economic Research, March 2020.

[4] Martin S Eichenbaum, Sergio Rebelo, and Mathias Trabandt. The macroeconomics of epidemics. Working Paper 26882, National Bureau of Economic Research, March 2020.

[5] Callum J Jones, Thomas Philippon, and Venky Venkateswaran. Optimal mitigation policies in a pandemic: Social distancing and working from home. Working Paper 26984, National Bureau of Economic Research, April 2020.

[6] Carlo A Favero. Why is covid-19 mortality in lombardy so high? evidence from the simulation of a seihcr model. Technical report, 2020.

[7] Marco Vergano, Guido Bertolini, Alberto Giannini, Giuseppe Gristina, Sergio Livigni, Giovanni Mistraletti, and Flavia Petrini. Raccomandazioni di etica clinica per l’ammissione e trattamenti intensivi e per la loro sospensione, in condizioni eccezionali di squilibrio tra necessitá e risore disponibili. Technical report, 2020.

[8] ISTAT. Decessi del 2020. dataset analitico con i decessi giornalieri., 2020. Data retrieved on May 4, 2020, https://www.istat.it/it/archivio/242055.

[9] ISTAT. Indicatori demografici. popolazione residente al 1° gennaio., 2020. Data retrieved on April 12, 2020, https://www.istat.it/it/popolazione-e-famiglie?dati.

[10] Gabriele Ciminelli and Sílvia Garcia-Mandico. COVID-19 in Italy: An Analysis of Death Registry Data. Journal of Public Health, 09 2020. fdaa165.

[11] Adam van Dijk, Don McGuinness, Elizabeth Rolland, and Kieran M Moore. Can telehealth ontario respiratory call volume be used as a proxy for emergency department respiratory visit surveillance by public health? Canadian Journal of Emergency Medicine, 10(1):18–24, 2008.

[12] Isaia Invernizzi. Coronavirus, chiamate al 118 giù del 90% ma l’attenzione resta alta - infografica, 2020. retrieved on October 10, 2020, https://www.ecodibergamo.it/.

[13] Regione Lombardia. Letti per struttura sanitaria di ricovero, 2020. Open Data Portal. retrieved on April 15, 2020, https://www.dati.lombardia.it/.

[14] ISTAT. Matrici di contiguità, distanza e pendoralismo, 2020. Data retrieved on October 10, 2020, https://www.istat.it/it/archivio/157423.

[15] Gabriele Ciminelli and Silvia Garcia-Mandico. Business shutdowns and covid-19 mortality. Preprint, 2020.

[16] Maria Sorbi. Un’ora di attesa per un’ambulanza. ora anche il 118 rischia il collasso, 2020. retrieved on May 13, 2020, https://www.ilgiornale.it.

[17] Regione Lombardia. Elenco rsa accreditate, 2020. Open Data Portal. retrieved on April 15, 2020, https://www.dati.lombardia.it/.

[18] ISTAT. Condizioni socio-economiche delle famiglie, 2018. Data retrieved on April 12, 2020, https://www.istat.it/it/archivio/190365.

[19] ISTAT. A misura di comune, 2020. Data retrieved on April 15, 2020, https://www.istat.it/it/archivio/220004.

[20] ISTAT. Dati ambientali nelle città. ambiente urbano, 2019. Data retrieved on April 12, 2020, https://www.istat.it/it/archivio/55771.

